# Persistence of viral RNA, widespread thrombosis and abnormal cellular syncytia are hallmarks of COVID-19 lung pathology

**DOI:** 10.1101/2020.06.22.20136358

**Authors:** Rossana Bussani, Edoardo Schneider, Lorena Zentilin, Chiara Collesi, Hashim Ali, Luca Braga, Ilaria Secco, Maria Concetta Volpe, Andrea Colliva, Fabrizio Zanconati, Giorgio Berlot, Furio Silvestri, Serena Zacchigna, Mauro Giacca

## Abstract

COVID-19 is a deadly pulmonary disease with unique clinical features. A thorough understanding of the molecular and histological correlates of the disease is still missing, especially because post-mortem analysis of COVID-19-affected organs has been so far scant and often anecdotical. Here we report the results of the systematic analysis of 41 consecutive post-mortem samples from individuals who died of COVID-19. We found that the disease is characterized by extensive alveolar damage and thrombosis of the lung micro- and macro-vasculature. Thrombi were in different stages of organization, consistent with an ongoing, endogenous thrombotic process. In all the analyzed samples, in situ RNA hybridization showed that pneumocytes and vascular endothelial cells had massive presence of viral RNA even at the later stages of the disease. An additional feature of the disease was the presence, in the vast majority of patients, of a large number of dysmorphic pneumocytes, often forming large syncytial elements, a consequence of the fusogenic activity of the viral Spike protein, detected with specific antibodies. Despite occasional presence of virus-positive cells in the heart, no overt signs of viral infection were detected in other organs, which showed common alterations compatible with prolonged hypoxia, multifocal organ disease or previous comorbidities. In summary, COVID-19 is a unique interstitial pneumonia with extensive lung thrombosis, long-term persistence of viral replication in pneumocytes and endothelial cells, along with the presence of infected cellular syncytia in the lung. We propose that several of the COVID-19 disease features are due to the persistence of virus-infected cells in the lungs of the infected individuals for the duration of the disease.

## INTRODUCTION

In December 2019, a novel strain of Coronavirus, named severe acute respiratory syndrome coronavirus-2 (SARS-CoV-2), was isolated from a few patients with COronaVIrus Disease 2019 (COVID-19) by the Chinese Center for Disease Control and Prevention and linked to the cluster of acute respiratory distress syndrome emerging in Wuhan, China ^1^. On 30 January 2020, the World Health Organization defined the outbreak of SARS-Cov-2 as an international, public health emergency. Clinical features of patients with COVID-19 indicate that infection with SARS-Cov-2 causes severe, and sometimes fatal, interstitial pneumonia similar to SARS-CoV infection, often requiring intensive care and oxygen therapy ^2^. Most patients present with fever, dry cough, dyspnea, and typical bilateral ground-glass opacities on chest CT scans.

Despite the rapid and worldwide spread of SARS-CoV-2, the pathology underlying the exceptional severity of COVID-19 in several individuals is still poorly understood. Increased amounts of proinflammatory cytokines (e.g, IL1B, IL6, IFNγ, IP10, and MCP1) have been documented in the blood of patients with extensive lung damage, suggestive of a cytokine storm condition associated with disease severity ^3,4^. After a first phase of viral replication, this inflammatory response was proposed to rapidly amplify even in the absence of significant viral load, resulting in systemic inflammation and eventually leading to multiple organ failure ^5^. Diffuse intravascular coagulation and large-vessel thrombosis have been linked to multisystem organ failure, likely associated to hypoxemia in multiple tissues ^6^, The type of lung pathology eventually causing this condition, however, has remained largely unexplained to date.

Other unknowns relate to the involvement of other organs in COVID-19. Besides indirect multi-organ injury, scattered reports have hinted at the possibility of direct injury caused by viral replication in brain, heart and kidney. Mechanistically, SARS-CoV-2 enters its target cells through the interaction of its Spike (S) envelope protein with the angiotensin-converting enzyme-2 (ACE-2) enzyme and upon activation by the TMPRSS2 plasma membrane protease ^7^. As both ACE2 and TMPRSS2 are broadly expressed in various tissues, including brain, heart and kidney ^8^ and in light of the similarity with SARS-CoV, which also interacts with ACE2 and had been found in both heart and brain of infected patients ^9,10^, it was proposed that infection with SARS-CoV-2 could also occur in these organs. A single case of encephalitis caused by SARS-CoV-2 was reported in Beijing, with evidence for the presence of viral RNA by genome sequencing in the cerebrospinal fluid ^11^. SARS-CoV-2, similar to other SARS coronaviruses, is detectable in the urine via PCR, indicating that kidney tubules are directly exposed to it ^12^. Electron microscope particles with SARS-CoV-2 morphology were shown in the kidney epithelium ^13^. However, despite these anecdotical reports, clear documentation of brain, kidney and heart direct damage caused by the virus is largely missing at this moment.

The systematic analysis of pathology of the lungs and other organs in COVID-19 patients would provide answers to these outstanding questions. However, evidence of disease by post-mortem analysis has been provided in only a few cases so far. Here were report a large and systematic study, performed in 41 consecutive patients who died of COVID-19 at the University Hospital in Trieste, Italy. We provide evidence that COVID-19 is a unique interstitial pneumonia in that it associates with extensive thrombosis of the lung macro- and micro-vasculature, the persistence of viral replication in the lung pneumocytes and endothelial cells for several weeks from diagnosis and the presence of abnormal cell syncytia in the lung. Other organs lacked overt signs of direct viral infection. We propose that several of the COVID-19 unique features are due to the persistence of abnormal, virus-infected cells in the lungs for prolonged periods of time.

## RESULTS

Here we report an extensive characterization of multiple post-mortem tissues from several organs of a large cohort of patients deceased of COVID-19. Of the 41 cases considered in our study (25 males and 16 females), 6 required intensive care, while 35 were hospitalized in either other hospital units (Geriatrics n = 12, Infectious Diseases n = 8, Pneumology n = 4, Clinical Medicine n = 2) or local Nurseries (n = 9) until death. We analyzed lung samples from all cases (n = 41). In addition, from several patients, we also collected and analyzed hearts (n = 30), kidneys (n = 23), livers (n = 23) and brains (n = 3). The average age of patients was 77 for males and 84 for females. Patient data are summarized in **Table I**. During hospitalization, all patients scored positive for SARS-CoV-2 on nasopharyngeal swab and presented symptoms and imaging data indicative of interstitial pneumonia related to COVID-19 disease.

### Lung damage is uniquely severe in COVID-19 patients requiring intensive care

Patients requiring intensive care (IC) with respiratory support were relatively younger (mean age: 69; age range: 43-82) compared to those hospitalized in other units (mean age: 84; age range: 47-97). All IC cases exhibited extremely severe lung damage. At macroscopic examination, lungs appeared congested (representative case in **Figure 1a**). In 4 cases, there was macroscopic appearance of thrombosis of large pulmonary vessels, often with multiple thrombi and in one case determining an extensive infarction in the right lobe (**Figure 1b** panels **a** and **b**). Histological analysis of all cases revealed gross destruction of the normal lung architecture (shown for representative patients in **Figure 2a** panels **a-c**), consistent with a condition of diffuse alveolar damage with edema, intra-alveolar fibrin deposition with hyaline membranes and hemorrhage. This was accompanied by massive occlusion of alveolar spaces due to cell delamination (panels **d-f**). Loss of cellular integrity was also confirmed by the presence of nuclear dust, indicative of ongoing cellular death (panel **g**). Numerous calcified areas were detected throughout the lung parenchyma of several patients (panel **h**). Quite unexpectedly, the inflammatory infiltrate was relatively modest in most cases (representative picture in **Figure 2b** panel **a**), mainly consisting of clusters of CD8+ lymphocytes (at two magnifications in panels **b** and **b’**) and relatively less abundant infiltration of CD4+ cells (panels **c** and **c’**), and the presence of macrophages (panels **d** and **d’**). In a few instances, perivascular lymphocytic caps, compatible with ongoing vasculitis, were observed (**Figure 2c**). These inflamed vessels were often characterized by endothelial shedding into the vascular lumen (**Figure 2d**). Remarkable vascular defects were observed in one IC individual, in whom numerous vessels of medium and small size were abnormal and stenotic due to massive deformation of their structure (**Suppl. Figure 1**). These vessels had an edematous wall, in which smooth muscle lost mutual contacts and endothelial cells appeared large, exfoliated and plumped. All three layers were largely inflamed, as revealed by the massive presence of immunoglobulins and HLA-DR-positive leukocytes.

**Figure 1.**
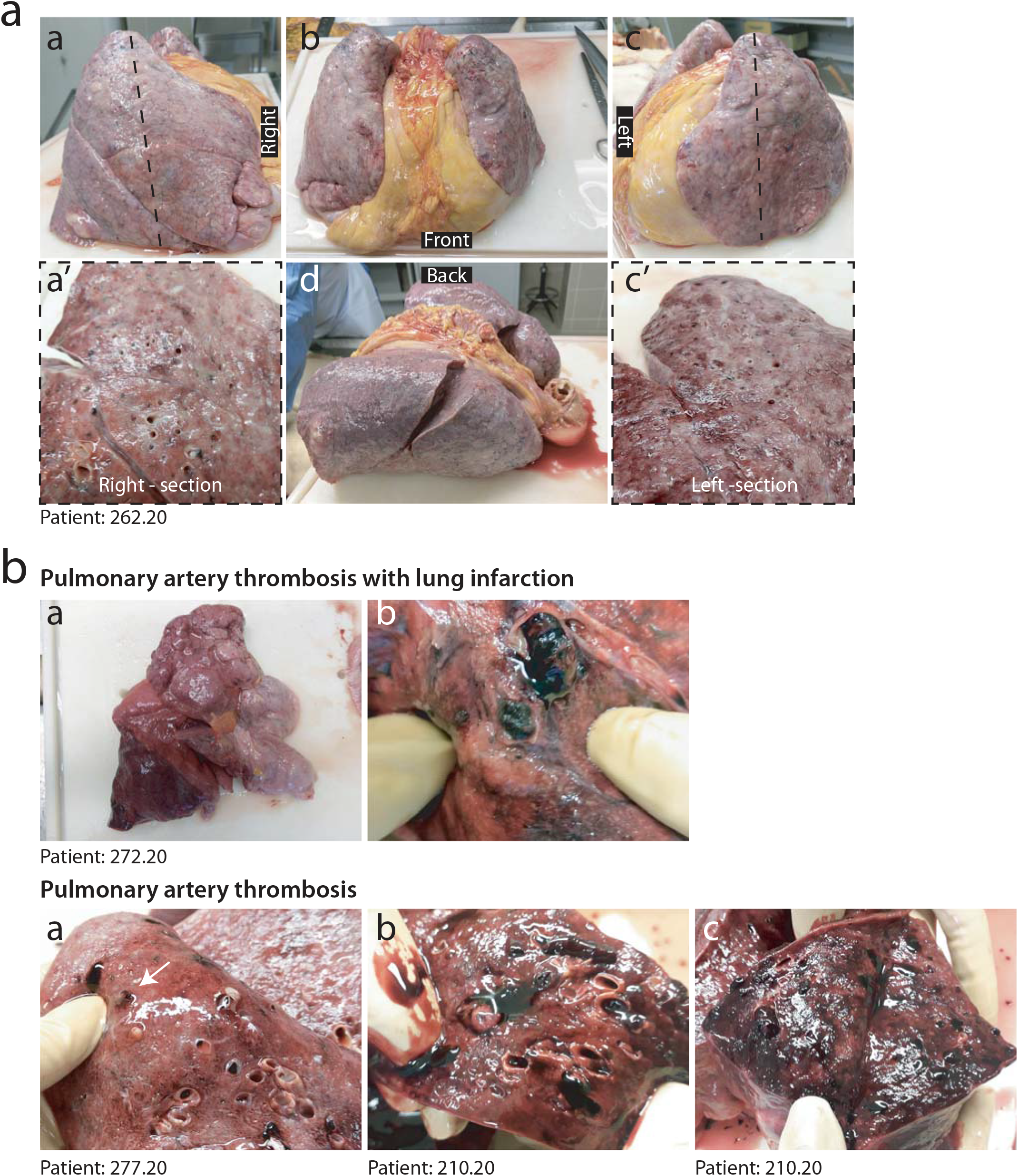
Macroscopic appearance of COVID lungs. **a**. Gross appearance of *en bloc* resection of heart and lungs from a severely affected IC patient, shown in its left, anterior, posterior and right views, as indicated (a-d). Lungs appeared congested and firm. Cut sections of both right and left lungs revealed tan-grey solid parenchyma with numerous hemorrhagic areas and loss of air spaces (a’, c’). **b**. A large thrombus in the inferior branch of the right pulmonary artery determined a large infarct in the right lung. Most lungs presented multiple thrombi, with loss of alveolar spaces and extensive hemorrhages (c-e).

**Figure 2.**
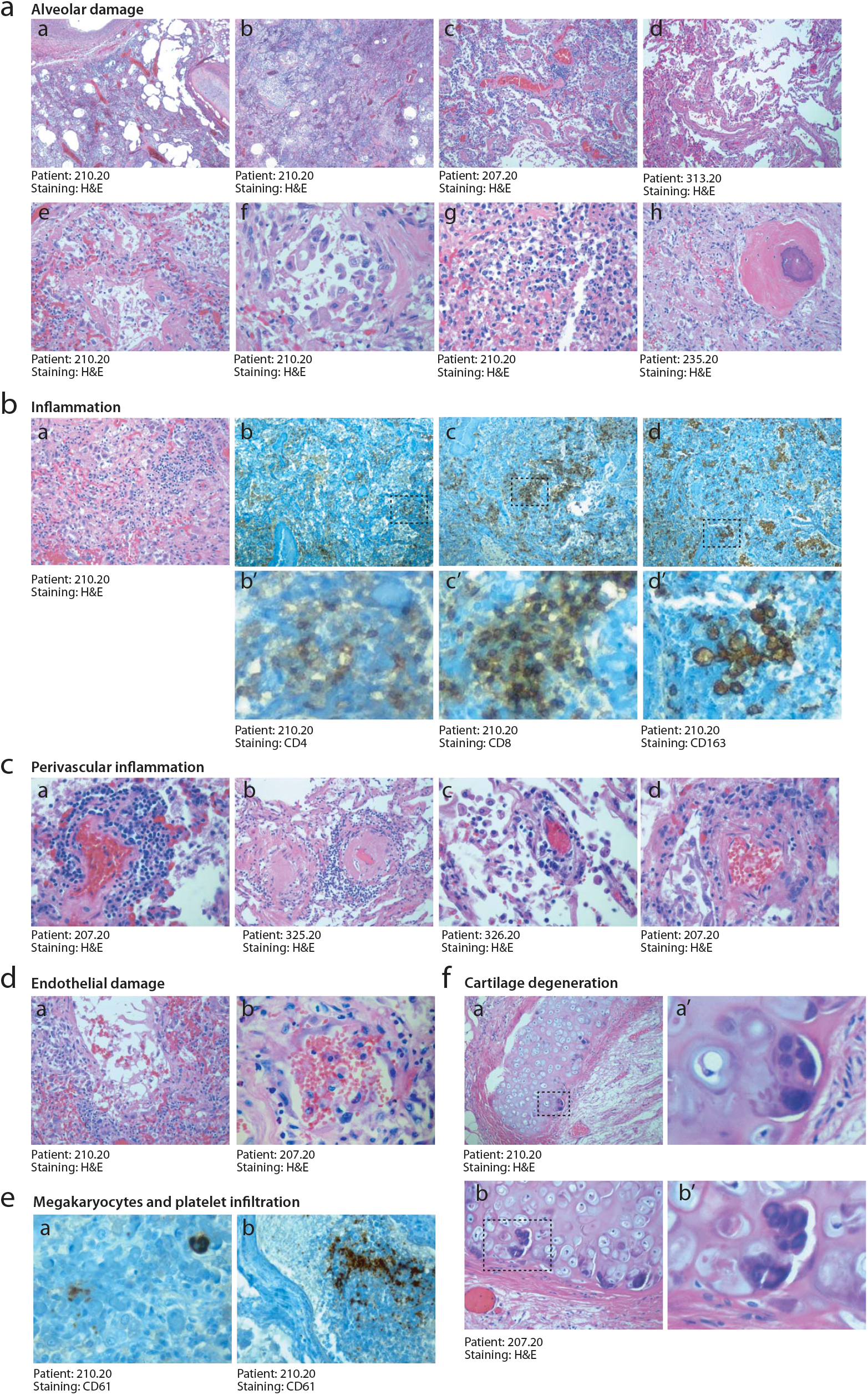
Histopathological evidence of massive cellular damage and inflammation in COVID-19 lungs. **a. Severe and diffuse alveolar damage**. Low magnification images (a, b, x2.5) show diffuse end-stage alveolar damage, with patchy inflammatory infiltrates (c, ×10), fibrinous exudate and hyaline membranes (c,d, ×10). Higher magnification images better demonstrate massive disruption of alveolar structures and hyaline membranes (e, x20), detachment of epithelial cells from the alveolar wall (f, x40), diffuse nuclear dust (g, x20) and limited foci of osseous metaplasia (h, ×10). **b. Moderate inflammatory infiltration**. In most patients, lungs were sparsely infiltrated by clusters of inflammatory cells (a), largely composed of CD4^+^ helper and CD8^+^ suppressor lymphocytes (low magnification images in b, c; high magnification images in b’, c’), as well as of CD163^+^ histiocytes (low magnification images in d; high magnification images in d’). In b-d’, nuclei were stained with hematoxylin. **c. Perivascular inflammation**. Numerous patients show clusters of inflammatory cells around small and medium size vessels. In all cases, perivascular lymphocytic cuffs were particularly evident around small arterioles, which had thick walls, often undergoing fibrinoid necrosis (a-d). **d. Severe and diffuse endothelial damage**. Medium and small vessels showed multiple vascular abnormalities, with massive liquefactive degeneration of endothelial cells (a) and their detachment from the vascular wall (b). **e. Infiltration by megakaryocytes and platelets**. Lungs affected by pulmonary thrombi appeared often infiltrated by sparse megakaryocytes (a) and by the presence of numerous platelets within vessels (b). Nuclei were stained with hematoxylin. **f. Cartilage abnormalities**. All IC patients showed unique alteration of the bronchial cartilage, with the appearance of multiple atypical chondrocytes, apparently multi-nucleated (low magnification images in a, b; high magnification images in a’, b’). In all panels, H&E: hematoxylin and eosin.

In several cases, the lung parenchyma was infiltrated by numerous megakaryocytes and platelet clusters were present in the thrombi occluding the lung vasculature (cf. later; **Figure 2e**). Finally, a peculiar and unexpected finding in all the analyzed individuals was the presence of atypical features of bronchial chondrocytes, which were often very large and dysmorphic (**Figure 2f** panels **a/a’** and **b/b’** at two magnifications).

### Vascular thrombosis and presence of atypical cells are hallmarks of COVID-19 lung pathology

Interstitial pneumonia from other causes tend to present with massive interstitial inflammation and diffuse alveolar damage (DAD), resulting in extensive pneumocyte loss ^14^. In contrast, lungs of COVID-19 patients showed relatively more modest inflammation but were characterized by two additional distinctive features. First, 5/6 IC patients (83%) and 16/35 non-IC patients (46%) showed the presence of massive lung vessel thrombosis. Thrombi were present in both the micro-and macro-vasculature. **Figure 3a** panels **a-h** show diffuse thrombosis of both small and large vessels in a series of analyzed patients. Thrombi were hetero-synchronous, showing different stages of organization, consistent with an ongoing thrombotic process endogenous to the lung. Sporadic lung thrombosis was also present in additional 8/35 of the non-IC patients, for a total of 29/41 (71%) of all the patients that were analyzed.

**Figure 3.**
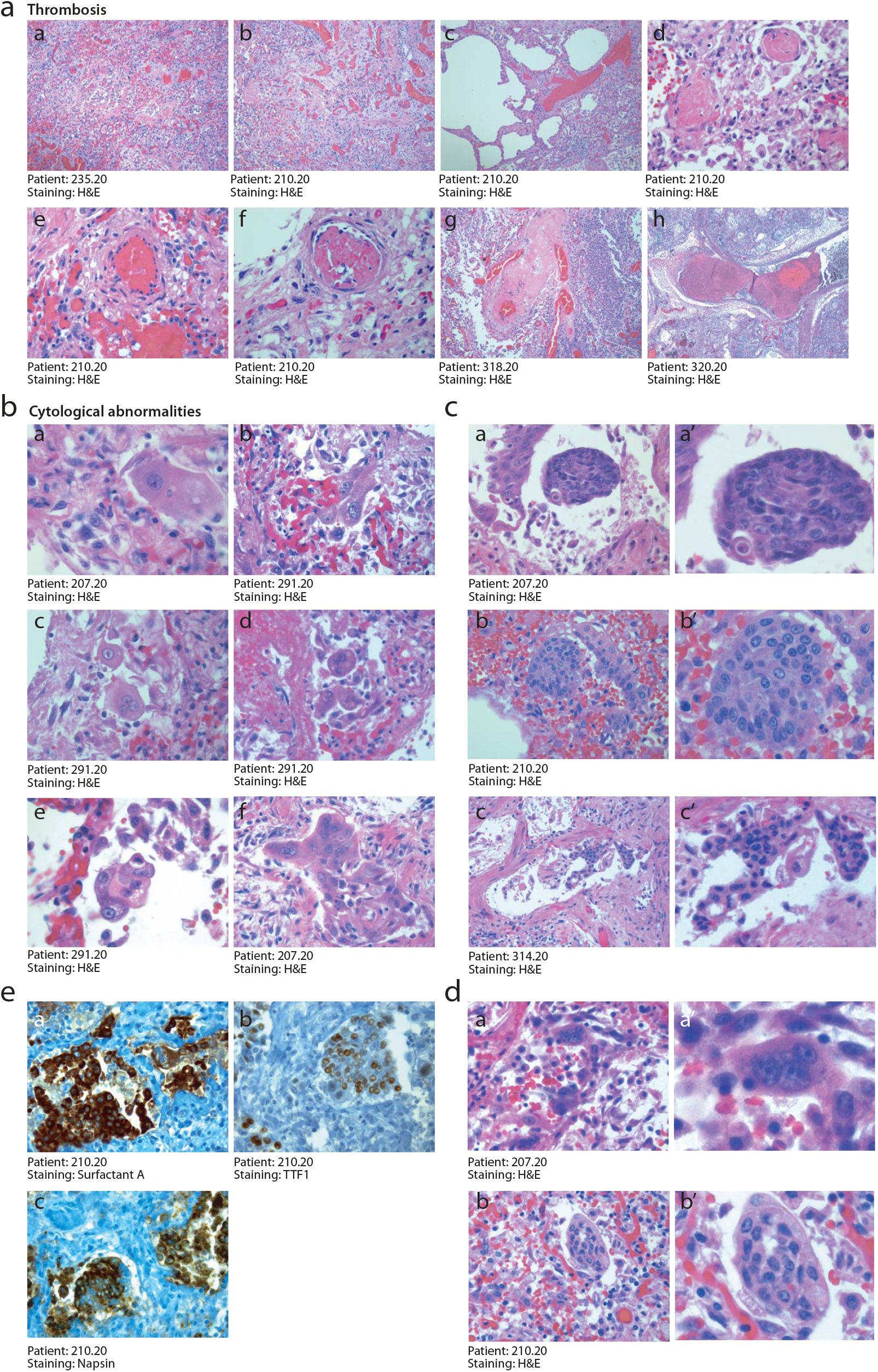
Histopathological evidence of massive thrombosis and unique cytological elements in COVID lungs. **a. Multiple thrombi in different stages of organization**. Consistent with the macroscopic detection of multiple thrombi in several pulmonary vessels, histological analysis revealed the presence of diffuse micro-thrombi (a, b, x2.5), which sometimes appeared rich in fibrin and poorly organized, consistent with their recent formation (c, ×10), but in most instances were largely organized (d, x20). High magnification images confirmed the co-existence of multiple thrombi in different stages of organization in the same lung (a recent thrombus and an organizing thrombus coexist in the lung of the same IC patient, shown in panels e and f, respectively, x20), as well as thrombotic lesions that have likely evolved structurally in multiple times, being composed of an older and a younger component (g, ×10; h, x2.5). **b. Unique cytological features of SARS-CoV-2**. A consistent and typical feature in COVID-19 lungs was the appearance of major cytological abnormalities, including the presence of giant cells (a, x63; b, x40), with a large cytoplasm, which often became bi-or multi-nucleated (c-f, x40). **c. Squamous metaplasia (pseudosyncytia)**. A common finding was the metaplasia of the alveolar epithelium, with a marked change in morphology of pneumocytes and their aggregation to form pseudosyncytia (a, b, c, 20x; a’, b’, c’, 63x). **d. Syncytia**. In addition to squamous metaplasia, true syncytial elements were observed in numerous lungs, showing large cytoplasm and nuclear aggregation (a, b, 20x; a’, b’, 63x). **e. Epithelial origin of syncytial elements**. The giant and multinucleated cells forming either pseudo-syncytia or real syncytia scored positive for the pneumocyte markers Surfactant-A (a, x20), TTF1 (b, x20) and Napsin (c, x20), indicative of their epithelial origin. Nuclei were stained with hematoxylin. In A-D, H&E: hematoxylin and eosin.

A second pathology hallmark of COVID-19 disease was the presence of anomalous epithelial cells. These were characterized by abnormally large cytoplasm and, very commonly, by the presence of bi-of multi-nucleation (**Figure 3b** panels **a-f**). In several cases, these cells were present in the alveolar spaces or jutted out into the neighboring, damaged vascular wall. In several instances, these cells clustered to form areas of squamous metaplasia (shown for three patients in **Figure 3c**, panels **a-c’** at two magnifications). Even more notably, the dysmorphic cells very often showed features of syncytia, characterized by several nuclei with an ample cytoplasm surrounded by a single plasma membrane (**Figure 3d** panels **a**-**b’** at two magnifications). These syncytia-forming, dysmorphic cells were bona fide pneumocytes, as they resulted positive for surfactant A and napsin, two pneumocyte-specific markers, and expressed the pneumocyte-specific transcription factor TTF1 in their nucleus (**Figure 3e** panels **a-c** respectively). Dysmorphic and syncytial pneumocytes were present and abundant in the lungs of 20 patients (50%), including all 6 patients requiring IC, and occasional in additional 16 patients (39%).

In addition to these COVID-19-specific alterations, the lungs of several patients, especially the elder in the non-IC group, showed signs of pre-existing morbidities, including emphysema and chronic obstructive pulmonary disease (**Table 1**).

**Table 1.** Selected clinical and histopathological features **Pneumonia severity score** 1, multiple micro-foci of modest inflammation 2, multiple micro-foci of moderate inflammation 3, diffuse moderate inflammation 4, diffuse severe inflammation 5, massive inflammation with loss of lung architecture * with *ab ingestis* component # with bacterial component **Abnormal pneumocytes** + rare ++ occasional +++ frequent § thromboembolic origin

| Case | Gender | Anticoagulant or antiaggregant therapy | Hosp. time (days) | Pneumonia severity | Abnormal pneumocytes | Lung thrombosis | Thrombosis in other organs |
| --- | --- | --- | --- | --- | --- | --- | --- |
| **Intensive Care (IC)** |  |  |  |  |  |  |  |
| 197.20 | M | Yes | 1 | 5 | +++ | Yes | No |
| 207.20 | F | No | 21 | 4 | +++ | No | No |
| 210.20 | M | No | 29 | 5 | +++ | Yes | No |
| 216.20 | M | No | 21 | 5* | +++ | Yes | No |
| 235.20 | M | Yes | 23 | 4 | +++ | Yes | No |
| 262.20 | M | Yes | 18 | 4 | +++ | Yes | No |
| **Other hospital units** |  |  |  |  |  |  |  |
| 165.20 | M | No | 14 | 2# | + | No | No |
| 241.20 | F | No | 3 | 4 | +++ | Yes | No |
| 257.20 | F | No | 10 | 4 | +++ | Yes | No |
| 272.20 | M | No | 30 | 5 | ++ | Yes | No |
| 276.20 | M | Yes | 18 | 5 | +++ | Yes | Yes§ |
| 277.20 | M | No | 8 | 3 | ++ | Yes | No |
| 285.20 | M | No | 26 | 3# | ++ | Yes | No |
| 288.20 | F | No | 28 | 4# | ++ | Yes | No |
| 301.20 | M | No | 8 | 2 | + | No | No |
| 302.20 | M | Yes | 60 | 2 | + | No | No |
| 303.20 | M | No | 17 | 4 | ++ | Yes | No |
| 304.20 | M | No | 9 | 3 | + | No | No |
| 305.20 | M | No | 14 | 3 | + | Yes | No |
| 306.20 | M | No | 24 | 3 | ++ | Yes | No |
| 307.20 | F | No | 7 | 4 | +++ | No | No |
| 308.20 | M | Yes | 6 | 4 | +++ | No | No |
| 309.20 | F | No | 5 | 4 | +++ | Yes | No |
| 310.20 | M | No | 8 | 5 | +++ | Yes | No |
| 311.20 | M | No | 15 | 4 | + | Yes | No |
| 312.20 | M | No | 14 | 3 | ++ | Yes | No |
| 313.20 | M | No | 17 | 2 | + | No | No |
| 314.20 | F | No | 24 | 3 | + | Yes | No |
| 315.20 | F | No | 28 | 3 | ++ | Yes | No |
| 316.20 | F | No | 7 | 2 | + | Yes | No |
| 317.20 | M | No | 4 | 2 | + | Yes | No |
| 318.20 | M | No | 2 | 4# | + | Yes | No |
| 319.20 | F | No | 6 | 4 | + | Yes | No |
| 320.20 | M | No | 13 | 5 | +++ | Yes | No |
| 321.20 | F | No | 14 | 1 | - | No | No |
| 322.20 | M | No | 27 | 5# | + | Yes | No |
| 323.20 | F | Yes | 27 | 3 | + | No | No |
| 324.20 | F | Yes | 14 | 3# | - | No | No |
| 325.20 | F | Yes | 48 | 1 | - | Yes | No |
| 326.20 | F | No | 3 | 4# | + | Yes | No |
| 327.20 | F | Yes | 19 | 2# | + | No | No |

### COVID-19 patients do not show overt signs of viral infection or ongoing inflammation in other organs

We analyzed the heart, liver and kidney in 6/6 and the brain in 3/6 IC patients. These samples presented common features of age-related tissue degeneration, including cardiac fibrosis, liver steatosis, kidney tubular micro-lithiasis, brain gliosis and neuronal loss. However, cellular morphology and tissue integrity were essentially preserved in all the analyzed samples (**Suppl. Figure 2a-c**). We did not detect clear signs of hepatitis, nephritis or encephalitis, such the presence of massive infiltration of inflammatory cells and/or with cytological alterations compatible with direct viral damage. Thrombosis was not detected in any of these organs.

Unspecific signs of age-related pathologies were also common in non-IC individuals. We carefully analyzed the myocardium in additional 19/35 of the non-IC patients. In all cases, the cardiac tissue showed variable extent of cardiomyocyte damage in both the right and left ventricles, consisting in myocardial fiber disarray, loss of sarcomeric material and cardiomyocyte anucleation (**Suppl. Figure 2d**), often accompanied by fibrosis and amyloid deposition. These pathological features are consistent with the presence of chronic and sustained hypoxia overimposed onto pre-existing damage. No evidence of inflammatory cell infiltration indicative of ongoing myocarditis was evident for any of these patients.

In 12/35 non-IC patients, we also harvested kidney tissue, which often exhibited tubular micro-lithiasis and epithelial necrosis. Also in these cases, no clear signs consistent with viral infection or acute inflammatory disease were observed.

### Detection of SARS-CoV-2 RNA reveals infection of lung pneumocytes and endothelial cells

We wondered whether the massive destruction of alveolar tissue, subverting the lung microarchitecture, accompanied by thrombosis and the presence of abnormal cells might results from persisting viral infection, despite several of these patient deceases after 30-40 days from first diagnosis. We designed two LNA-modified RNA probes for the SARS-CoV-2 genome, hybridizing to the positive strand viral RNA. Negative hybridization controls were samples from normal individuals deceased for other causes in 2018 (representative sample in **Figure 4a** panel **a**), positive controls were samples hybridized with a probe for the ubiquitous U6 RNA, showing nuclear localization, as per our previous experience ^15^; (**Figure 4a** panel **b**).

**Figure 4.**
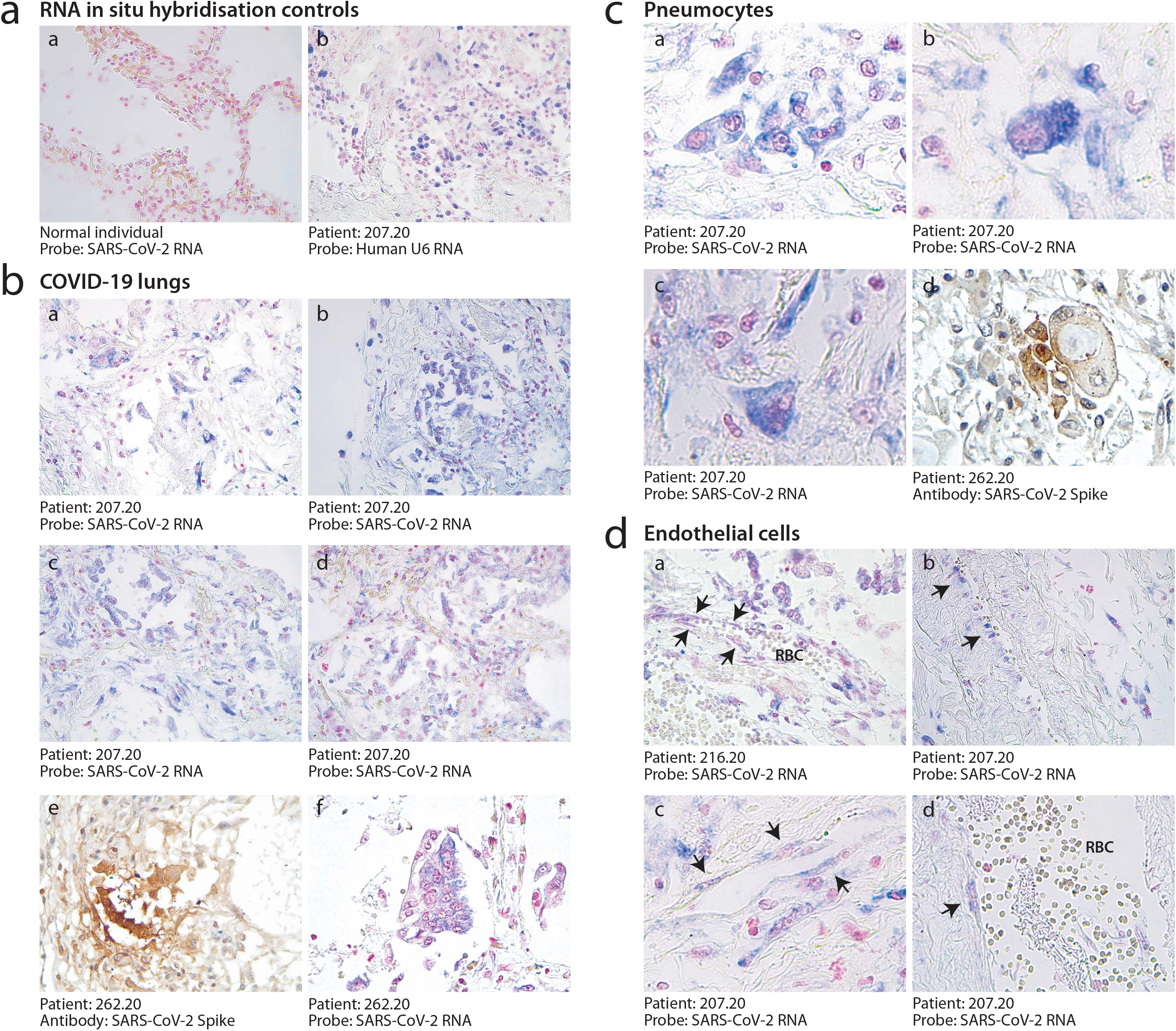
Presence of SARS-CoV-2 RNA in lungs of COVID-19 patients. **a. Controls for ISH**. The probe specific for SARS-CoV-2 RNA did not develop any signal in autoptic lung samples harvested in 2018, when the virus was not circulating in humans (a, ×10). ISH for human ribosomal U6 showed nuclear staining in all cells, consistent with its ubiquitous expression (b, ×10). **b. Detection of SARS-CoV-2 in COVID-19 lungs**. SARS-CoV-2 ISH revealed a specific and diffuse signal in the atypical cells (a-d, ×10; e, x20, f, × 40) detected in lungs of COVID-19 patients. Cuboidal pneumocytes positive for the expression of the Spike protein (e). Pseudosyncytia were positive for viral RNA (f). **c Presence of SARS-CoV-2 in pneumocytes**. Intense SARS-CoV-2 RNA signal was mainly detected in giant and multinucleated cells (a-c, x63), which were identified as pneumocytes (Figure 2E). Some of these cells also expressed the Spike protein (S) of SARS-CoV-2 (d, x63). **d. Presence of SARS-CoV-2 RNA in endothelial cells**. SARS-CoV-2 signal was present in elongated cells, localized along the luminal side of vessel wall, compatible with their endothelial origin (a,b,×10; c, d, x40). Nuclei were stained with nuclear fast red for ISH and with hematoxylin for IHC.

Lung samples from 10 patients were analyzed by in situ hybridization with an equimolar mixture of the 2 SARS-CoV-2 probes. This study revealed the massive presence of virus-infected cells in these individuals (representative pictures shown in **Figure 4a** panels **a-d**). The viral genome was also expressed, as concluded from positivity for viral Spike protein expression (panel **e**). The cells undergoing squamous metaplasia resulted infected (panel **f**). Similarly, the abnormally large and polynucleated pneumocytes detected by histology and immunostaining were also virtually all infected, showing a large cytoplasm with intense staining for the probe (**Figure 4a** panels **a-c**). Several cells, including those showing gross cytological abnormalities, not only showed the presence of the viral RNA but also expressed viral proteins (shown for the Spike protein in **Figure 4a** panel **d**). Of note, several of these dysmorphic, Spike-positive cells showed highly expanded cytoplasms with the presence of large inclusion bodies.

Several endothelial cells lining the pulmonary vessels also showed clear evidence of viral RNA presence (representative images from two individuals shown in **Figure 4d** panels **a-d**). Small vessels positive for the presence of the viral genome were also occasionally found in the heart (**Suppl. Figure 2d** panel **f**), along with the sporadic presence of viral RNA also in other cell types (panels **g** and **h**).

## DISCUSSION

Here we report the results of the analysis of the so far largest series of 41 consecutive patients who died from COVID-19. Our findings add new important information on the unique pathology suffered by these individuals and on the extent of viral replication in their lungs.

In the patients who died after prolonged intensive care, the extent of alveolar damage was massive, as previous studies have already revealed in more limited cohorts ^16,17^. Damage was not dissimilar from that of patients succumbing from acute respiratory distress syndrome (ARDS) secondary to other conditions, including detachment of alveolar pneumocytes, deposition of hyaline membranes, marked interstitial edema with advanced fibrotic organization and immune cell infiltration. Besides the lung, however, in our series of post-mortem analyses we could not detect overt sign of possible viral replication in other organs, including brain, liver, heart and kidney. Most of the patients who succumbed to COVID-19 had an advanced age and their organs showed variegate signs of age-related comorbidities, none of which was specifically be reminiscent of viral infection. This does not exclude, however, that infection of a very few cells in these organs would still be a cause for disease. Indeed, despite the absence of histological evidence of overt inflammation, we could occasionally detect small vessels and other isolated cells positive for the viral RNA in the heart.

Our *in situ* RNA hybridization for the detection of the SARS-CoV-2 genome univocally indicates that the alterations in the lung are concomitant with persistent viral infection. RNA-positive pneumocytes were present in large numbers in the lungs of 10/10 tested individuals. There has been a suggestion that COVID-19 follows a biphasic course, with an initial phase characterized by viral replication, also detectable by positivity at the qualitative, RT-PCR-based swab test, followed by second phase in which viral replication might become less relevant, characterized by hyperinflammation. While the clinical manifestations of the disease remain different while disease progresses, our data challenge the notion that viral replication has ceased in patients with advanced disease. On the contrary, the presence of abundant cytoplasmic RNA signals in the lungs of the deceased patients after 30-40 days from diagnosis is clearly suggestive of ongoing replication and postulate a role for infection in sustaining both disease phases. Thus, despite swab tests can become negative over time, infection of lung pneumocytes can persist over the entire course of the disease.

In addition to persistent viral infection, two additional hallmarks appear to characterize COVID-19 pathology. First, the presence of massive lung thrombosis. This was evident at macroscopic examination in 5/6 patients from IC Unit and in 31/41 patients in the whole cohort analyzed at the histological level. Fibrin deposition and thrombosis affected both small and large pulmonary vessels and showed clear signs of heterochronicity, with several fresh thrombi neighboring thrombi in advanced stage of fibrotic organization. This is indicative of an endogenous pro-thrombotic process ongoing in the lungs as opposed as being the lung the target for an embolic dissemination event. In terms of defining the underlying cause for thrombosis, it is tempting to associate the widespread presence of SARS-CoV-2-infected cells with the induction of a pro-thrombotic state. Consistent with this possibility, we found several endothelial cells lining small and larger vessels infected with the virus, also confirming previous, more anecdotic evidence ^18^. Infection of endothelial cells is rendered possible by the known expression of the viral ACE2 receptor in these cells ^19^. The presence of lung thrombosis has earlier been associated clinically with COVID-19 disease and recently reported histologically in a smaller group of 7 post-mortem analyses ^16^ and in another series of patients from Northern Italy (https://doi.org/10.1101/2020.04.19.20054262). These findings are also consistent with biochemical evidence of activation of the coagulation process, in particular high level of circulating D-dimer ^20,21^. Whether these thrombi can resolve in response to anti-coagulant therapy is still matter of debate ^22^. In one of our IC patients, who showed transient clinical improvement upon treatment with high dose of anti-coagulants, we could observe an old, organized thrombus detached from the arterial wall, consistent with re-canalization of the vessel **(Suppl. Figure 1D)**.

Besides persistent viral infection and lung thrombosis, our study indicates that a third hallmark of COVID-19 disease is the presence of dysmorphic cells in the lung, which were abundant and easily detected in all the analyzed patients. As concluded from the expression of pneumocyte markers, these cells were abnormal pneumocytes, commonly with an enlarged cytoplasm, inclusion bodies and very often characterized by bi-or tri-nucleation. Most strikingly, in several occasions and in virtually all patients, multinucleated syncytia were also apparent. While squamous metaplasia is often found in lungs from other ARVD cases, as per our common experience and also reported by others for COVID-19 ^23,24^, these syncytia, which were clearly morphologically different, appeared to be characteristic of COVID-19 disease.

The presence of these syncytial cells is likely to be the direct consequence of the expression of the Spike (S) protein of SARS-CoV2. Different from SARS-CoV, the type I fusogenic protein of SARS-CoV-2 present in the infectious virions becomes activated directly at the level of the plasma membrane by cellular proteases, most notably TMPRSS2 ^7^. This feature permits envelope-plasma membrane fusion and virion internalization without requiring prior virion endocytosis. As a consequence of this characteristic, the presence of the S protein on the membrane of SARS-CoV-2-infected cells determines fusion of these cells with other neighboring cells expressing ACE2 and TMPRSS2. In vitro expression of the S protein in permissive cells causes the formation of large syncytia, which are highly reminiscent of those observed in vivo, an effect that is largely inapparent with the S protein of SARS-CoV (data not shown).

In light of the persistence of virus-infected cells in the lungs of infected individuals and the presence of these syncytial elements we propose that several of the peculiar characteristics that set COVID-19 apart from other interstitial pneumonias are not attributable to pneumocyte death as a consequence viral replication, but to the persistence of virus-infected-S-expressing cells in the lungs of the infected individuals. Whether these features, in the individuals who survive severe COVID-19 disease, result in permanent damage and lead to long-term consequences on lung structure and function is an outstanding question, which will require future clinical and pathological investigation.

## Data Availability

All data concerning this manuscript will be rendered available

## Acknowledgments

We are very grateful to Giacomo Molinari, Joseph Abu Nasr, Katia Di Vito, Sonia De Luca, Marzia Pavlovich, Paola Suligoi, Ida Rosano, Gabriella Slatich and Lorena Ulcigrai for excellent technical support in autopsies, histology and immunohistochemistry. The authors are grateful to Ajay Shah for critical comments and suggestions. Part of this work was supported by a King’s Together Rapid COVID-19 Call grant from King’s College London. MG is supported by Programme Grant RG/19/11/34633 and the King’s College London Centre of Research Excellence grant RE/18/2/34213 from the British Heart Foundation.

## METHODS

### Case cohort

Histological analysis was performed by expert technicians and pathologists at the Pathology Unit of Trieste University Hospital, which has reached a remarkably high autopsy rate over more than one century (approximately 50 % of hospital deaths). In most cases, full body autopsy was performed, which allowed us to analyze lung, brain, heart and kidney tissues. The same pathologists analyzed all samples considered in this study, excluding operator-dependent biases. Clinical data, including age, gender, known co-morbidities and therapies were collected and listed in **Table 1**. This study was approved by the competent Joint Ethical Committee of the Regione Friuli Venezia Giulia.

### Histology and immunohistochemistry

At least five samples were collected from each organ, selecting representative areas at macroscopic examination. Samples were fixed in 10% formalin for at least 50 hours and then embedded in paraffin. One-micron sections were processed for the following histological and immunohistochemistry (IHC) staining: hematoxylin-eosin, Azan Mallory, Alcian Blue PAS (ABPAS), colloidal iron, CD4, CD8, CD163, CD61, Napsin A, TTF1, *α*-SMA, CD31 (all from Roche), HLA-DR (from Neomarkers), Surfactant B (from Thermo), and against the SARS-CoV-2 Spike protein (from GeneTex). Antigen retrieval techniques and antibody pretreatment were performed according to the manufacturer’s specifications. Images were acquired using a Leica DM2700 M light microscope.

### In situ hybridization

In situ hybridization (ISH) was performed using locked nucleic acid (LNA) probes for U6 snRNA ^15^ and SARS-CoV-2 RNA, designed to target the sense strand of ORF1ab and Spike regions of the viral genome. Scrambled sequences were used as control. Experiments were performed using a dedicated ISH kit for Formalin-fixed paraffin-embedded (FFPE) tissues (Qiagen) according to the manufacturer’s protocol. Briefly, FFPE tissue slides were deparaffinized in xylene, treated with proteinase-K (15 µg/ml) for 5 min at 37°C and incubated with either SARS-CoV-2 (40 nM) or U6 probes (2 nM) for one hour at 54°C in a hybridizer. After washing with SSC buffer, the presence of SARS-CoV-2 RNA was detected using an anti-DIG alkaline phosphatase (AP) antibody (1:500) (Roche Diagnostics) supplemented with sheep serum (Jackson Immunoresearch) and bovine serum albumin (BSA). Hybridization was detected by adding NBT-BCIP substrate (Roche Diagnostics). Nuclei were counterstained with nuclear fast red.

---

**Supplementary Figure 1.**
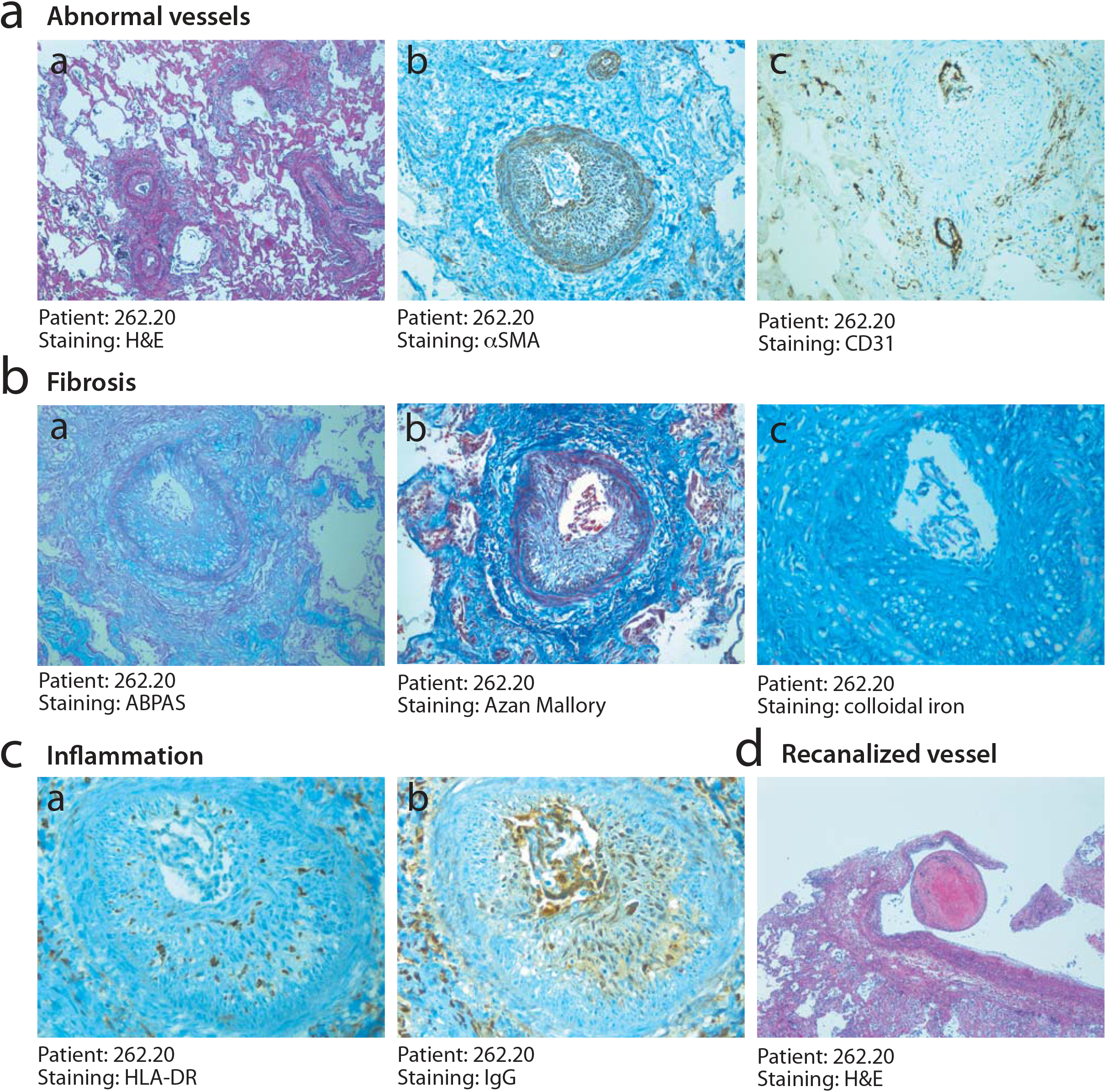
Severe vessel disease in one IC patient. **a. Abnormal vessel structure**. Multiple medium-size vessels appeared stenotic with abnormal wall structure (a, x2.5), due to the presence of massive edema that caused disaggregation of smooth muscle cells (b, ×10) and endothelial cell detachment (c, ×10). H&E: hematoxylin and eosin. **b Fibrosis within the arterial wall**. The medial layer of the arterial wall included fibrotic tissue of recent formation, as shown by Alcian Blue PAS (a, ×10), Azan Mallory (b, ×10) and colloidal iron (c, x20) staining. Nuclei were stained with hematoxylin. **c. Inflammation of the media and intima**. Massive inflammation in the medial and intimal layer of the arterial wall was documented by HLA-DR (a, x20) and immunoglobulin (IgG, b, x20) staining. Nuclei were stained with hematoxylin. **d. Vessel recanalization after anticoagulant therapy**. An old, well organized thrombus appeared detached from the vascular wall, possibly indicating recanalization of a previously occluded artery (x2.5). H&E: hematoxylin and eosin.

**Supplementary Figure 2.**
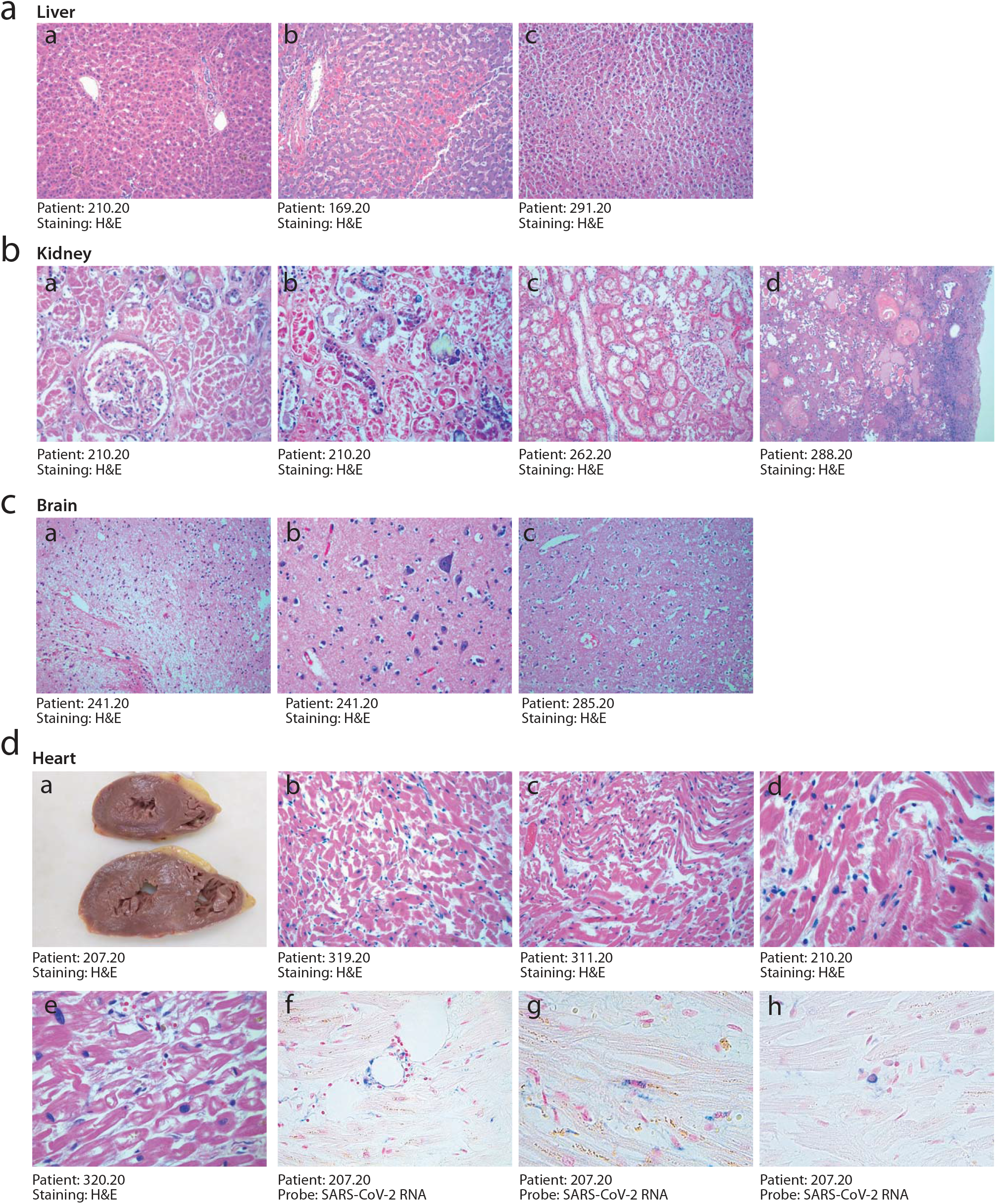
Absence of histopathological signs compatible with acute viral infections in other organs of COVID-19 patients. **a. Liver**. Liver parenchyma of patients affected by the most severe pneumonia either had a normal appearance (a, c, ×10) or showed unspecific signs, such as hemorrhagic congestion (b, ×10), in the absence of overt inflammation. **b. Kidney**. In most patients, kidneys showed preserved parenchymal structure (a, x20), often in the presence of either micro-(b, x20) or macro-lithiasis (d, ×10), and hemorrhagic congestion (c, ×10). **c. Brain**. All brains presented signs of neurodegeneration consistent with the advanced age of the patients, which included neuronal loss and gliosis (a, c, ×10) and vascular rarefaction (b, x20). **d. Heart**. Macroscopic examination of heart cuts in COVID-19 patients revealed hypoperfused cardiac muscles (a). Histological analysis revealed loss of apoptotic cardiomyocytes with nuclear loss, perinuclear halos and wavy myocells (b, c, x20; d, e, x40). ISH with a SARS-CoV-2 probe revealed the occasional presence of small vessels (f, x40) and other isolated cells (g and h, 40x) positive for the viral genome. Presence of these cells was sporadic (1/10 hearts analyzed). Nuclei in f-h were stained with nuclear fast red. In all panels, H&E: hematoxylin and eosin.

**Supplementary Table 1.** Clinical and histopathological features Units IC, Intensive Care; GE, Geriatrics; IN, Infectious Diseases; PN, Pneumology; NU, Nursery; IM, Internal Medicine Pneumonia severity score: 1, multiple micro-foci of modest inflammation 2, multiple micro-foci of moderate inflammation 3, diffuse moderate inflammation

| Case | Gender | Co-morbidities | Therapy | Hosp. time (days) | Pneumonia severity | Abnormal pneumocytes | Lung thrombosis | Thrombosis in other organs |
| --- | --- | --- | --- | --- | --- | --- | --- | --- |
| **Cases from Intensive Care (IC)** |  |  |  |  |  |  |  |  |
| 197.20 | M | Dementia, Parkinson disease, Ischemic cardiomyopathy, Hypertension | Amiodarone, Bisoprolol, Diuretics, Aspirin, Warfarin, Thyroxine | 1 | 5 | +++ | Yes | No |
| 207.20 | F | Aortic valve replacement, cardiac pacemaker | Aspirin, Colchicine, Ibuprofen, Acenocumarol | 21 | 4 | +++ | No | No |
| 210.20 | M | Ischemic cardiomyopathy | Hydroxychloroquine | 29 | 5 | +++ | Yes | No |
| 216.20 | M | Pulmonary hypertension, diabetes | Diltiazem, Tadalafil, Aspirin, Acetylcysteine, Statins | 21 | 5* | +++ | Yes | No |
| 235.20 | M | Prostate cancer, pyonephrosis, previous pulmonary embolism | Heparin | 23 | 4 | +++ | Yes | No |
| 262.20 | M | Diabetes, Bladder cancer, Hypercholesterolemia | Insulin | 18 | 4 | +++ | Yes | No |

Supplementary Table 1. Clinical and histopathological features
|  |  |  |  |  |  |  |  |  |
| --- | --- | --- | --- | --- | --- | --- | --- | --- |
| 165.20 | M | Aortic valve replacement, cardiac pacemaker, Hypertension, Lung cancer | Warfarin, Bisoprolol, Statins, Sartans, Salmeterol | 14 | 2# | + | No | No |
| 241.20 | F | Dementia, Stroke | Aspirin, Diazepam | 3 | 4 | +++ | Yes | No |
| 257.20 | F | Previous lung embolism, COPD, lung, uterus and kidney cancer, diverticulitis, PAOD | Apixaban, Amiodarone, Bisoprolol, Spironolactone, Diuretics | 10 | 4 | +++ | Yes | No |
| 272.20 | M | Seizures, urosepsis, atrial fibrillation, hypertension, encephalopathy | Carbamazepin, Lacosamide, Prednisone, Bisoprolol, Amdiodarone, Aspirin, Triazolam, Ramipril | 30 | 5 | ++ | Yes | No |
| 276.20 | M | Leriche syndrome with lower limb amputation, diabetes, atrial fibrillation, hypertension | Insulin, Clopidogrel, Rivaroxaban, Diuretics, Doxazosin, Sartans, Alfuzosin | 18 | 5 | +++ | Yes | Yes\$ |
| 277.20 | M | Diabetes, Hypertension, chronic kidney disease | Insulin, Sartans | 8 | 3 | ++ | Yes | No |
| 285.20 | M | Disseminated lung cancer, ischemic cardiomyopathy, diabetes, hypertension | Statins, Sartans, Bisoprolol, Metformin, Glimepiride, Aspirin | 26 | 3# | ++ | Yes | No |
| 288.20 | F | Chronic kidney disease, ischemic cardiomyopathy, atrial fibrillation, previous lung embolism, nephrolithiasis, pyonephrosis | Isosorbide, Diuretics, Spironolactone, Bisoprolol, Aspirin, Warfarin, Tamsulosin, Vitamin D | 28 | 4# | ++ | Yes | No |
| 301.20 | M | Chronic kidney disease, psychosis, gastric cancer | Haloperidol, Triazolam, Folate, Diuretics, Calcium | 8 | 2 | + | No | No |
| 302.20 | M | Psychosis, dementia, antiphospholipid syndrome, hypertension | Aspirin, Heparin | 60 | 2 | + | No | No |
| 303.20 | M | Hypokinetic syndrome, tibia and fibula fracture, diabetes, hypertension, prostate cancer | Insulin, Bisoprolol, Sartans | 17 | 4 | ++ | Yes | No |
| 304.20 | M | Hypertensive cardiomyopathy, stroke | Furosemide, Apixaban, Statins, Ramipril, Dutasteride | 9 | 3 | + | No | No |
| 305.20 | M | Diabetes, heart failure, chronic kidney disease, urinary tract infection, hypertension | Insulin, Nitrates, Bisoprolol, Sartans | 14 | 3 | + | Yes | No |
| 306.20 | M | COPD, heart failure | Bisoprolol, Apixaban, Furosemide, Salbutamol | 24 | 3 | ++ | Yes | No |
| 307.20 | F | Bronchiectasis, hypertension, diabetes | Insulin, Salbutamol, Diltiazem, Amiodarone | 7 | 4 | +++ | No | No |
| 308.20 | M | Dysphagia, ab ingestis pneumonia, acute kidney disease, decubitus, vascular dementia, sigma cancer | Aspirin, Heparin | 6 | 4 | +++ | No | No |
| 309.20 | F | Lower limbs ulcers, heart failure, pancreatic cancer, bronchiectasis, atrial fibrillation | Warfarin, Diuretics, Bisoprolol, Sprinolactone, Fentanil | 5 | 4 | +++ | Yes | No |
| 310.20 | M | Dementia, hypertension | Sartans, Tamsulosin | 8 | 5 | +++ | Yes | No |
| 311.20 | M | Dementia, prostate hyperplasia, cholelithiasis, asthma | Lormetazepam, Prednisone, Salbutamol | 15 | 4 | + | Yes | No |
| 312.20 | M | Heart failure, mitral valve replacement, hypertension, hypertension, brain haemorrhage | Diuretics, Ramipril, Finasteride, Tamsulosin | 14 | 3 | ++ | Yes | No |
| 313.20 | M | Chronic kidney disease, cardiac amyloidosis, atrial fibrillation, hypertension, diabetes, ischemic cardiomyopathy | Bisoprolol, Diuretics, Calcium | 17 | 2 | + | No | No |
| 314.20 | F | Parkinson disease, breast cancer, facet joint arthrosis | Levo-dopa, Aspirin, Exerestane, Trazodone, Methylprednisolone | 24 | 3 | + | Yes | No |
| 315.20 | F | Cervical cancer, peritoneal carcinomatosis | Perindopril/Indapamide, Aspirin | 28 | 3 | ++ | Yes | No |
| 316.20 | F | Multiple fractures, osteoporosis | Heparin, Lorazepam, Vitamin D | 7 | 2 | + | Yes | No |
| 317.20 | M | Dementia, heart failure | Bisoprolol, Furosemide, Allopurinol, Statins, Warfarin | 4 | 2 | + | Yes | No |
| 318.20 | M | Dementia, urinary tract infection | Aspirin, Rivastigmine | 2 | 4# | + | Yes | No |
| 319.20 | F | Dementia, diabetes | Vitamin D, Tiroxin, Omega-3 fatty acids, Quetiapine, Statins, Sitagliptin, Insulin | 6 | 4 | + | Yes | No |
| 320.20 | M | Ataxia, seizures, nephrolithiasis, tracheostomy | Valproic acid, Olanzapine, Salbutamol, Lacosamide | 13 | 5 | +++ | Yes | No |
| 321.20 | F | Vertebral collapse, atrial fibrillation, paraparesis, syncopes, hypertension | Bisoprolol, Targin, Paracetamol | 14 | 1 | - | No | No |
| 322.20 | M | COPD, heart failure, diabetes, stroke | Bisoprolol, Clopidogrel, Diuretics, Aldactone, Insulin | 27 | 5# | + | Yes | No |
| 323.20 | F | Gastric ulcer, anemia, fibrothorax, knee endoprosthesis, femur fracture, hypertension | Alprazolam, Alendronic acid, Rivaroxaban, Bisoprolol, Digoxin | 27 | 3 | + | No | No |
| 324.20 | F | Diverticulosis, colon cancer, dementia, breast cancer, gastric cancer | Furosemide, Ramipril, Aldactone, Lorazepam, Aspirin, Clopidogrel | 14 | 3# | - | No | No |
| 325.20 | F | Heart failure, diabetes, dementia, femur fracture | Bisoprolol, Ramipril, Diuretics, Spironolactone, Metformin, Warfarin | 48 | 1 | - | Yes | No |
| 326.20 | F | Dementia, meningioma | Carbamazepine | 3 | 4# | + | Yes | No |
| 327.20 | F | Femur fracture, dementia | Heparin | 19 | 2# | + | No | No |

## Notes

### Competing Interest Statement

The authors have declared no competing interest.

### Funding Statement

Part of this work was supported by a Kings Together Rapid COVID-19 Call grant from Kings College London. MG is supported by Programme Grant RG/19/11/34633 and the Kings College London Centre of Research Excellence grant RE/18/2/34213 from the British Heart Foundation. The authors or their institutions have not received payment from a third party of any aspect of the submitted work

### Author Declarations

This study was approved by the competent Joint Ethical Committee of the Regione Friuli Venezia Giulia, Italy

